# COVID infection rates, clinical outcomes, and racial/ethnic and gender disparities before and after Omicron emerged in the US

**DOI:** 10.1101/2022.02.21.22271300

**Authors:** Lindsey Wang, Nathan A. Berger, David C. Kaelber, Pamela B. Davis, Nora D. Volkow, Rong Xu

## Abstract

**Background:** SARS-CoV-2 infections and hospitalizations are rising in the US and other countries after the emergence of the Omicron variant. Currently, data on infection rates, severity and racial/ethnic and gender disparities from Omicron in the US is limited.

**Method:** We performed a retrospective cohort study of a large, geographically diverse database of patient electronic health records (EHRs) in the US. The study population comprised 881,473 patients who contracted SARS-CoV-2 infection for the first time between 9/1/2021-1/16/2022, including 147,964 patients infected when Omicron predominated (Omicron cohort), 633,581 when Delta predominated (Delta cohort) and another 99,928 infected when the Delta predominated but just before the Omicron variant was detected in the US (Delta-2 cohort). We examined monthly incidence rates of COVID-19 infections stratified by age groups, gender, race and ethnicity, compared severe clinical outcomes including emergency department (ED) visits, hospitalizations, intensive care unit (ICU) admissions, and mechanical ventilation use between propensity-score matched Omicron and Delta cohorts stratified by age groups (0-4, 5-17, 18-64 and ≥ 65 years), and examined racial/ethnic and gender differences in severe clinical outcomes.

**Findings:** Among 147,964 infected patients in the Omicron cohort (average age: 39.1 years), 56.7% were female, 2.4% Asian, 21.1% Black, 6.2% Hispanic, and 51.8% White. The monthly incidence rate of COVID infections (new cases per 1000 persons per day) was 0.5-0.7 when Delta predominated, and rapidly increased to 3.8-5.2 when Omicron predominated. In January 2022, the infection rate was highest in children under 5 years (11.0) among all age groups, higher in Black than in White patients (14.0 vs. 3.8), and higher in Hispanic than in non-Hispanic patients (8.9 vs. 3.1). After propensity-score matching for demographics, socio-economic determinants of health, comorbidities and medications, risks for severe clinical outcomes in the Omicron cohort were significantly lower than in the Delta cohort: ED visits: 10.2% vs. 14.6% (risk ratio or RR: 0.70 [0.68-0.71]); hospitalizations: 2.6% vs. 4.4% (RR: 0.58 [0.55-0.60]); ICU admissions: 0.47% vs. 1.00% (RR: 0.47 [0.43-0.51]); mechanical ventilation: 0.08% vs. 0.3% (RR: 0.25 [0.20-0.31]). Similar reduction in disease severity was observed for all age groups. There were significant racial/ethnic and gender disparities in severe clinical outcomes in the Omicron cohort, with Black, Hispanic patients having more ED visits and ICU admissions than White and non-Hispanic patients, respectively and women had fewer hospitalization and ICU admission than men.

**Interpretation:** The incidence rate of COVID infection during the omicron predominant period (prevalence >92%) was 6-8 times higher than during the Delta predominant period that preceded it consistent with greater infectivity. The incidence rate was highest among those less than 5 years of age, and in Black and Hispanic patients. COVID infections occurring when the Omicron predominated were associated with significantly less frequent severe outcomes than in matched patients when the Delta variant predominated. There were significant racial, ethnic and gender disparities in severe clinical outcomes, with Black and Hispanic patients and men disproportionally impacted.

## Introduction

SARS-CoV-2 infections and hospitalizations are rising in the US and other countries after the emergence of the Omicron variant(1). Reports from South Africa(2), Scotland(3), and England(4) showed lower rates of hospitalization following Omicron infection compared with the Delta variant infection. Currently, data on disease infection rates, severity, and racial/ethnic and gender disparity from Omicron in both pediatric and adult populations in the US is lacking. Here we compared incidence rates of COVID-19 infections, severe clinical outcomes including ED visits, hospitalizations, ICU admissions, and mechanical ventilation use, and racial and ethnic disparities of infection rates and severe outcomes before and after Omicron emergence in the US, through a retrospective study of a large, geographically diverse database of patient electronic health records (EHRs) in the US. Outcomes in pediatric patients, adults and older adults were examined separately.

## METHODS

### Study population

This study used the TriNetX Analytics network platform that contains de-identified EHR data of 90 million unique patients from 63 health care organizations in both inpatient and outpatient settings across the US(5). TriNetX Analytics provides web-based secure access to patient EHR data from hospitals, primary care, and specialty treatment providers, covering diverse geographic locations, age groups, racial and ethnic groups, income levels, and insurance types. Although the data are fully de-identified, end-users can use built-in statistical functions to perform patient-level data analysis, including cohort selection, propensity-score matching, time trend analysis, outcome research, among others. Because this study only queried statistics of de-identified patient records through web-applications and did not involve retrieval, storage, collection, use, or transmittal of individually identifiable data, Institutional Review Board approval and informed consent was not needed or sought.

The study population comprised three cohorts of patients with first time SARS-CoV-2 infections: (a) Omicron cohort (n = 147,964) – contracted first SARS-CoV-2 infection between 12/26/2021– 1/16/2022. The CDC’s national genomic surveillance program reports that Omicron accounted for 92%-99% of all circulating virus variants in the US during the two week period of 12/26/2021–1/20/2022(6); (b) Delta cohort (n = 633,581) – contracted first SARS-CoV-2 infection between 9/1/2021–11/15/2021 when Delta was the predominant variant (99.0%)(6); (c) Delta-2 cohort (n = 99,928) – contracted first SARS-CoV-2 infection between 11/16/2021– 11/30/2021, immediately before the Omicron variant was detected in the US and when Delta was the predominant variant (99.0%)(6). This second Delta cohort was created to control for later time periods and shorter window of infection.

The status of SARS-CoV-2 infection was based on the ICD-10 diagnosis code of “COVID-19” (U07.1) or lab-test confirmed presence of “SARS coronavirus 2 and related RNA” (9088). The status of adverse clinical outcomes was based on the Current Procedural Terminology (CPT) relevant codes for ED visits (“Emergency Department Visits”, code 1013711), hospitalizations (“hospital inpatient services”, code: 013659), ICU admissions (“Critical Care Services”, code: 1013729), and mechanical ventilation use (“Respiratory ventilation”, codes: 5A1935Z, 5A1945Z, 5A1955Z, 5A09357, 5A09457, 5A09557).

### Statistical analysis

We examined monthly incidence rate of SARS-CoV-2 infections (new cases per 1,000 person per day) between 9/1/1021-1/20/2022 among patients who had no prior infections, stratified by age groups (0-4, 5-17, 18-64 and ≥65 years old), race and ethnic groups (Black, White, Hispanic and Non-Hispanic) and gender.

We tested whether severe clinical outcomes among patients in the Omicron cohort differed from those in the Delta cohort. The two cohorts were propensity-score matched (1:1 using a nearest neighbor greedy matching with a caliper of 0.25 times the standard deviation) for demographics (age, gender, race/ethnicity); adverse socioeconomic determinants of health (assessed by ICD-10 codes “Z55-Z65” for “Persons with potential health hazards related to socioeconomic and psychosocial circumstances”) that include employment, housing, education, and economic circumstances; comorbidities relevant to COVID-19 risks or outcomes(7,8) including hypertension, heart diseases, cerebrovascular diseases, cancer, obesity, type 2 diabetes, chronic respiratory diseases, chronic kidney diseases, liver diseases, HIV infection, dementia, substance use disorders, depression and anxiety (assessed by ICD-10 codes); behavioral factors (tobacco smoking, alcohol drinking) (assessed by one or more encounter based on ICD-10 codes); COVID-19-related medications(9) (assessed by RxNorm codes); and vaccination status documented in patient EHRs (assessed by CPT or RxNorm codes).

Severe clinical outcomes between the propensity-score matched Omicron and Delta cohorts and between propensity-matched Delta-2 and Delta cohorts was assessed based on the percentage of ED visits, hospitalizations, ICU admissions and need for mechanical ventilation in the 3-day time-window that followed from the first day of SARS-CoV-2 infection. Overall risk, risk ratios, and 95% confidence interval (CI) were calculated. Racial, ethnic and gender disparities were examined by comparing clinical outcomes (ED visits, hospitalization, ICU admission) in the 3-day time-window that followed from the first day of SARS-CoV-2 infection between matched Black and White patients, between matched Hispanic and non-Hispanic patients, and between women and men, for the Omicron, Delta, and Delta-2 cohort respectively. Mechanical ventilation was not examined due to limited sample sizes. Race, ethnicity and gender stratified cohorts (Black vs White, Hispanic vs. Non-Hispanic, women vs. men) were propensity-score matched for other demographics, socioeconomic factors, COVID-19-reated health conditions, and medications, and documented vaccination status. Overall risk, risk ratios, and 95% confidence interval (CI) were calculated.

All statistical tests (incidence rates and outcome comparisons) were conducted on 1/20/2022 within the TriNetX Analytics Platform with significance set at p-value < 0.05 (two-sided).

## Results

### Patient characteristics

The study population comprised 881,473 patients who contracted SARS-CoV-2 infection for the first time between 9/1/2021-1/16/2022, including 147,964 in the Omicron cohort, 633,581 in the Delta cohort, and another 99,928 in the Delta-2 cohort. The characteristics of the Omicron and Delta cohort before and after propensity-score matching are shown in **Table 1**. Compared to the Delta cohort, the Omicron cohort were older (average age: 39.1 vs 36.4 years), differed in gender, racial and ethnic compositions, had fewer comorbidities and adverse social determinants of health and less vaccination. After propensity-score matching for variates in the table, the differences between the two cohorts decreased with the Omicron cohort having more comorbidities and similar vaccination status.

**Table 1.** Characteristics of the Omicron cohort and Delta cohort before and after propensity matching. Omicron cohort –patients who first contracted SARS-CoV-2 infection between 12/26/2021–1/16/2022 when Omicron was predominant and accounted for 92%-99.5% of all variants circulating in the US. Delta cohort –patients who first contracted SARS-CoV-2 infection between 9/1/2021–11/15/2021 when Delta was predominant and accounted for 99% of all variants circulating in the US. Race and ethnicity as recorded in the TriNetX EHR database were included because they have been associated with both infection risk and severe outcomes of SARS-CoV-2 infections. P-value – significance between the two cohorts based on two-tailed two-proportion z-test conducted within the TriNetX Network. * Vaccination rates only include those in the TriNetX EHR database but does not capture all vaccinations outside the health care organizations.

|  | Before Matching |  |  | After Matching |  |  |
| --- | --- | --- | --- | --- | --- | --- |
|  | Omicron cohort | Delta Cohort | P-value | Omicron cohort | Delta cohort | P-value |
| **Total number of patients** | 147,964 | 633,581 |  | 147,107 | 147,107 |  |
| **Age (years, mean±SD)** | 39.1 ± 23.4 | 36.4 ± 25.2 | < .001 | 39.1 ± 23.4 | 38.9 ± 23.9 | 0.11 |
| **Sex (%)** |  |  |  |  |  |  |
| Female | 56.7 | 54.1 | < .001 | 56.7 | 56.4 | 0.07 |
| Male | 43.2 | 45.8 | < .001 | 43.2 | 43.6 | 0.07 |
| **Ethnicity (%)** |  |  |  |  |  |  |
| Hispanic/Latinx | 6.2 | 8.6 | < .001 | 6.1 | 6.3 | 0.13 |
| Not Hispanic/Latinx | 51.8 | 60.5 | < .001 | 51.7 | 50.9 | < .001 |
| Unknown | 42.0 | 30.8 | < .001 | 42.2 | 42.9 | < .001 |
| **Race (%)** |  |  |  |  |  |  |
| Asian | 2.4 | 2.4 | 0.06 | 2.4 | 2.5 | 0.16 |
| Black | 21.1 | 15.8 | < .001 | 21.1 | 21.1 | 0.71 |
| White | 60.0 | 62.5 | < .001 | 60.0 | 59.3 | < .001 |
| Unknown | 15.9 | 18.8 | < .001 | 15.9 | 16.5 | < .001 |
| **Adverse Social determinants of health (%)** | 2.1 | 3.1 | < .001 | 2.2 | 2.0 | 0.002 |
| **Comorbidities (%)** |  |  |  |  |  |  |
| Hypertension | 17.8 | 18.3 | < .001 | 17.8 | 16.9 | < .001 |
| Heart diseases | 3.9 | 4.9 | < .001 | 3.9 | 3.7 | 0.002 |
| Cerebrovascular diseases | 2.8 | 3.6 | < .001 | 2.8 | 2.6 | < .001 |
| Obesity | 11.5 | 11.5 | 0.93 | 11.5 | 10.7 | < .001 |
| Type 2 diabetes | 6.9 | 7.6 | < .001 | 6.9 | 6.3 | < .001 |
| Cancers | 12.6 | 13.7 | < .001 | 12.6 | 11.9 | < .001 |
| Chronic respiratory diseases | 11.1 | 13.3 | < .001 | 11.2 | 10.6 | < .001 |
| Liver diseases | 2.9 | 3.7 | < .001 | 2.9 | 2.6 | < .001 |
| Chronic kidney disease | 2.9 | 3.4 | < .001 | 2.9 | 2.6 | 0.001 |
| Blood disorders involving immune mechanisms | 11.1 | 13.0 | < .001 | 11.1 | 10.4 | < .001 |
| HIV infection | 0.17 | 0.23 | < .001 | 0.17 | 0.16 | 0.41 |
| Dementia | 0.4 | 0.6 | < .001 | 0.4 | 0.4 | 0.21 |
| Substance use disorders | 7.7 | 9.5 | < .001 | 7.7 | 7.4 | < .001 |
| Depression | 8.0 | 8.7 | < .001 | 8.0 | 7.4 | < .001 |
| Anxiety | 14.0 | 13.9 | 0.26 | 14.0 | 13.0 | < .001 |
| Smoking | 2.3 | 2.4 | < .001 | 2.3 | 2.1 | 0.05 |
| Alcohol abuse | 1.0 | 1.5 | < .001 | 1.0 | 1.0 | 0.20 |
| **Organ Transplant (%)** | 0.4 | 0.5 | < .001 | 0.4 | 0.3 | 0.09 |
| **COVID-19 therapeutics (%)** |  |  |  |  |  |  |
| Dexamethasone | 15.2 | 16.9 | < .001 | 15.2 | 14.2 | < .001 |
| Remdesivir | 0.41 | 0.37 | 0.05 | 0.41 | 0.40 | 0.58 |
| Hydrocortisone | 6.4 | 7.8 | < .001 | 6.4 | 6.0 | < .001 |
| Ibuprofen | 21.4 | 24.1 | < .001 | 21.4 | 20.7 | < .001 |
| Prednisone | 12.8 | 11.2 | < .001 | 12.8 | 12.0 | < .001 |
| Methylprednisolone | 17.9 | 13.3 | < .001 | 17.9 | 16.8 | < .001 |
| Prednisolone | 3.1 | 4.8 | < .001 | 3.1 | 3.2 | 0.29 |
| Naproxen | 7.5 | 6.2 | < .001 | 7.5 | 7.0 | < .001 |
| Fluoxetine | 2.5 | 2.5 | 0.96 | 2.5 | 2.2 | < .001 |
| Fluvoxamine | 0.09 | 0.08 | 0.18 | 0.09 | 0.08 | 0.34 |
| Tocilizumab | 0.06 | 0.08 | 0.05 | 0.06 | 0.05 | 0.16 |
| Casirivimab/ Imdevimab | 0.2 | 0.6 | < .001 | 0.2 | 0.2 | 0.70 |
| Ritonavir/ Lopinavir | 0.07 | 0.05 | 0.04 | 0.07 | 0.05 | 0.07 |
| **\*Vaccination (% of age ≥ 5 years)** | 7.7 | 13.6 | < .001 | 7.7 | 7.7 | 0.25 |

### Monthly incidence rates of first time SARS-CoV-2 infections between 9/1/2021-1/20/2022

The monthly incidence rate of SARS-CoV-2 infections (measured by new cases per 1000 persons per day) among patients without prior infections was mostly stable between September and November 2021 when Delta was the predominant strain: 0.6, 0.54, 0.7 for September, October, and November, respectively. The incidence rates rapidly increased to 1.3 in the first half of December 2021, coincident with the emergence of Omicron in the US. It reached 3.8 in the second half of December 2021 and 5.2 in January 2022, indicating that infections with Omicron have not plateaued during this period (**Figure 1a**). The infection rate was the highest in children under 5 years and reached 11.0 in January 2022, which is 6-9 times of the infection rate during the Delta period between September and November and 3 times of infection rate during the Omicron emergence period in the early December. Overall infection rates decreased with increasing age. Accordingly, patients over 65 years old had the lowest incidence rate (**Figure 1a**). Infection rates were higher in men than in women for both Delta and Omicron periods though differences were small and remained constant (**Figure 1b**).

**Figure 1.**
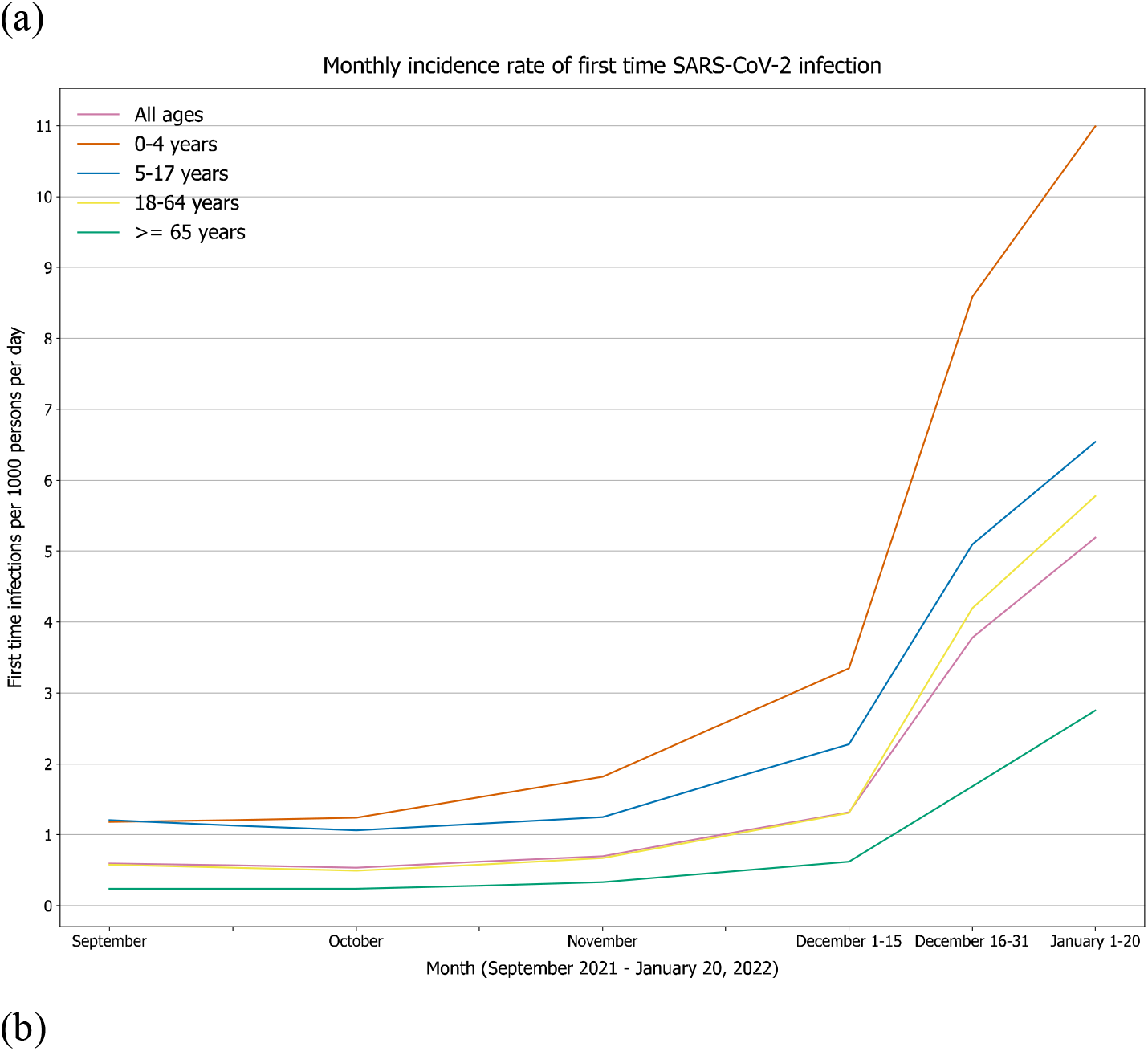
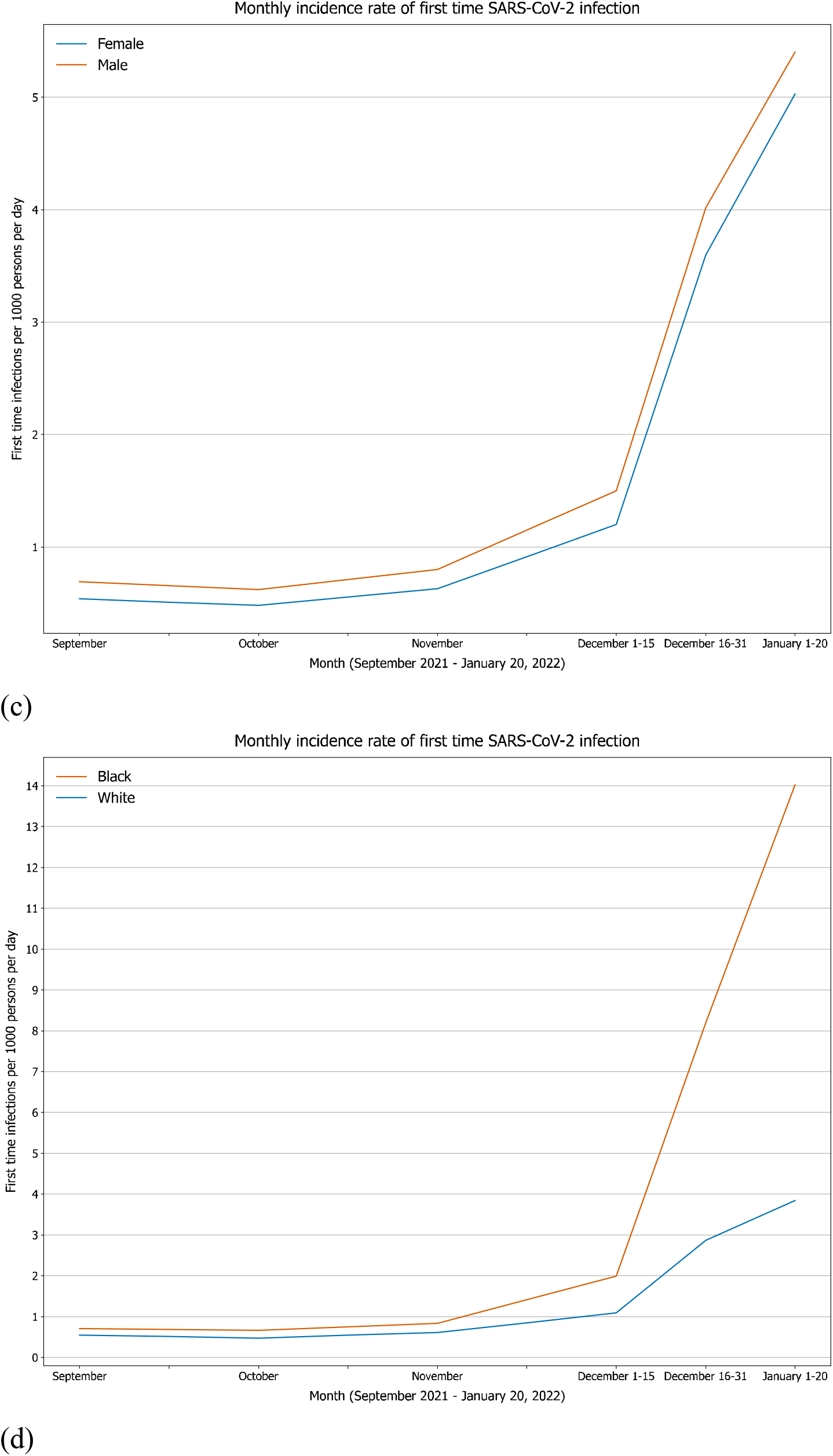
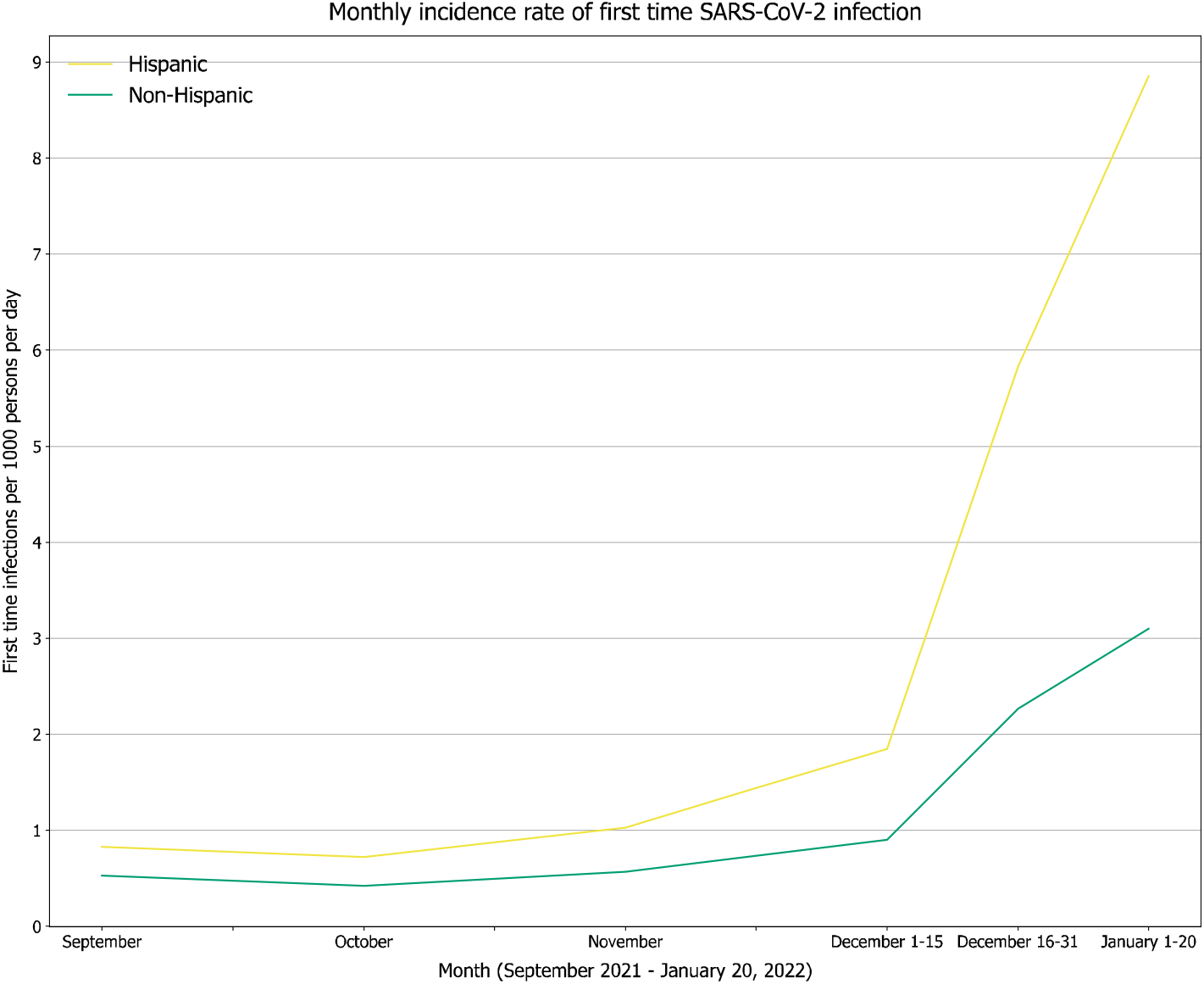
Monthly incidence rates of first-time SARS-CoV-2 infections (measured by new cases per 1000 person per day) between 9/1/2021-1/20/2022 in patients stratified by (a) age groups (0-4 years, 5-17 years, 18-64 years, and >65 years), (b) gender (Female, Male), (c) race (Black, White), and (d) ethnicity (Hispanic, Non-Hispanic).

Significant differences were observed between Black and White patients in both the Delta period and Omicron period, and the racial disparity was widened during the Omicron predominance period (**Figure 1c**). Between 12/26/2021-1/20/2022, the incidence rate among Black patients was 8.2-14.0, which were 2.9-3.7 times of the rates in White patients (P <0.001). During the Delta predominance period, the incidence rate among Black patients was 1.3-1.4 times of that in White patients. Significant differences were observed between Hispanic and non-Hispanic patients and the ethnic disparity was widened during the Omicron predominance period (**Figure 1d**). Between 12/26/2021-1/20/2022, the incidence rate among Hispanic patients was 5.8-8.9, which were 2.6-2.9 times of the rates in non-Hispanic patients (P <0.001). During the Delta predominance period, the incidence rate among Hispanic patients was 1.6-1.8 times of that in non-Hispanic patients. Significant racial and ethnic disparities in infection rate were consistent across all age groups during the Omicron predominance period (data not shown).

### Fewer severe clinical outcomes in the Omicron than in the matched Delta cohort

The risks for severe clinical outcomes in the 3-day time-window following initial SARS-CoV-2 infection in the Omicron cohort (n=147,107) were significantly lower than in the matched Delta cohort: ED visits: 10.2% vs. 14.6% (risk ratio or RR: 0.70 [0.68-0.71]); hospitalizations: 2.6% vs. 4.4% (RR: 0.58 [0.55-0.60]); ICU admissions: 0.47% vs. 1.00% (RR: 0.47 [0.43-0.51]); mechanical ventilation: 0.08% vs. 0.3% (RR: 0.25 [0.20-0.31]) (**Figure 2**, top). In contrast, the comparison between the matched Delta-2 and the Delta cohort showed higher prevalence of severe outcomes in the Delta-2 cohort compared to the Delta cohort presumably reflecting waning immunity from vaccination (**Figure 2**, bottom). Reductions in disease severity in the Omicron compared to the Delta cohort were significant for all age groups except for the 5-17 year group in whom the reductions in mechanical ventilation only achieved trend level significance. (**Figure 3**).

**Figure 2.**
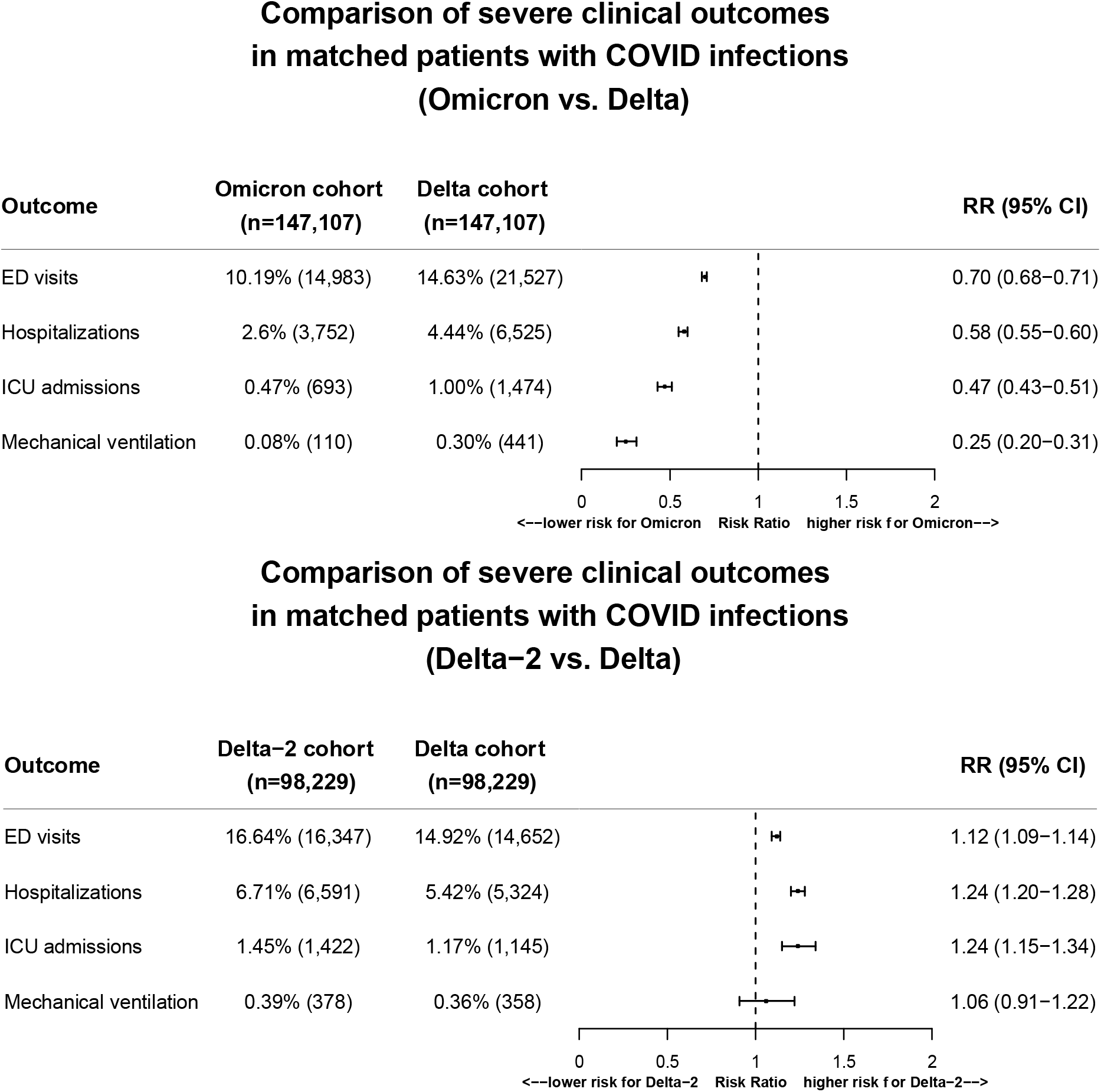
Comparison of severity of clinical outcomes including ED visits, hospitalizations, ICU admissions, and mechanical ventilation use in the 3-day time-window that followed from the first day of SARS-CoV-2 infection between matched Omicron and Delta cohorts (Top panel) and between matched Delta2 and Delta cohorts (Bottom panel). Omicron cohort – patients who first contracted SARS-CoV-2 infection between 12/26/2021–1/16/2022 when Omicron was predominant and accounted for 92-99.5% of all variants circulating in the US. Delta cohort – patients who first contracted SARS-CoV-2 infection between 9/1/2021–11/15/2021 when Delta was predominant and accounted for >99% of all variants circulating in the US. Delta-2 cohort – patients who first contracted SARS-CoV-2 infection between 11/16/2021–11/30/2021, right before the Omicron emergence in the US and when Delta accounted for >99% of all variants. Cohorts were propensity-score matched for demographics (actual age at index event of COVID infection, gender, race/ethnicity), socioeconomic factors, COVID-19-reated health conditions and medications, and documented vaccination status.

**Figure 3.**
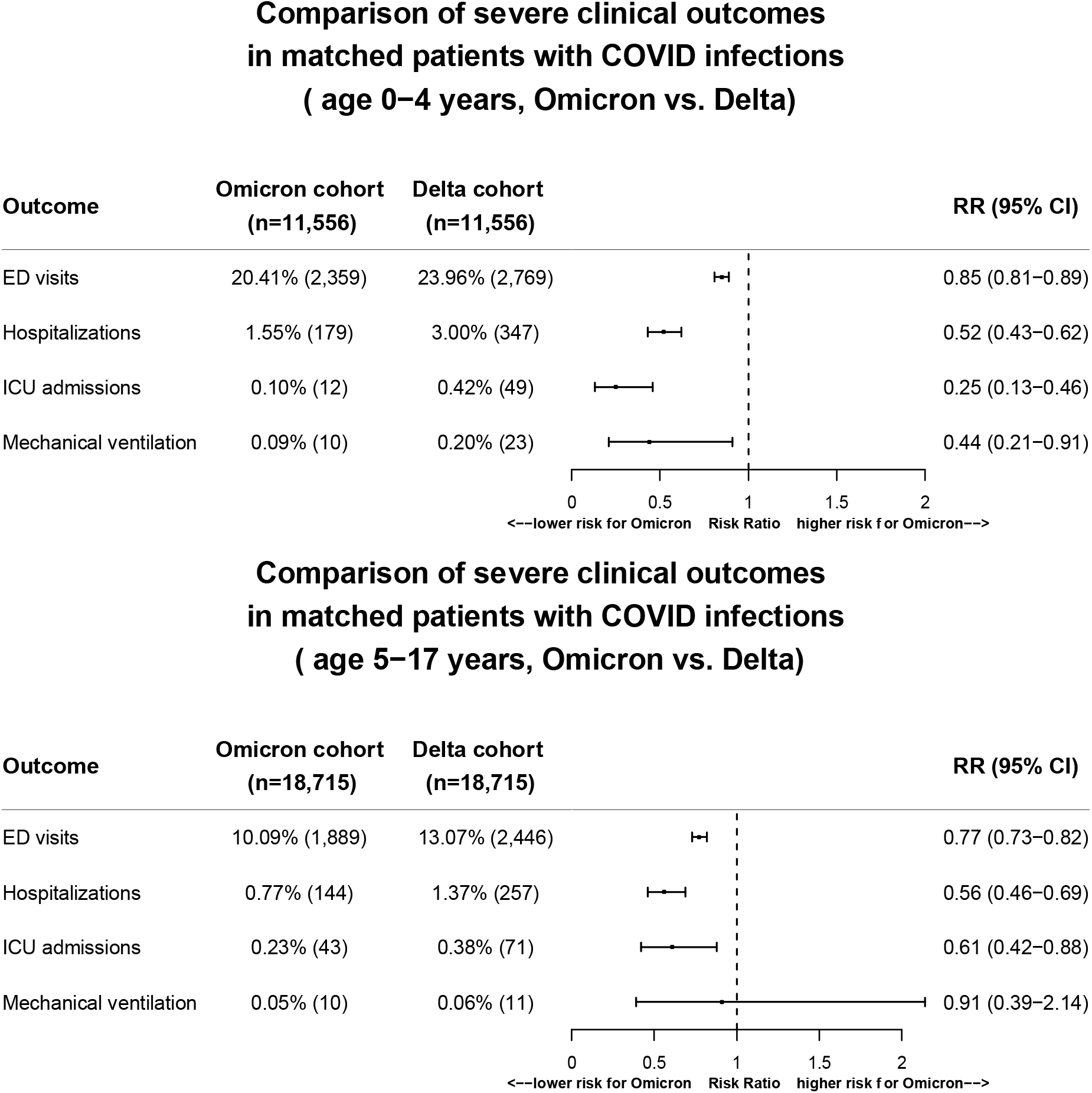
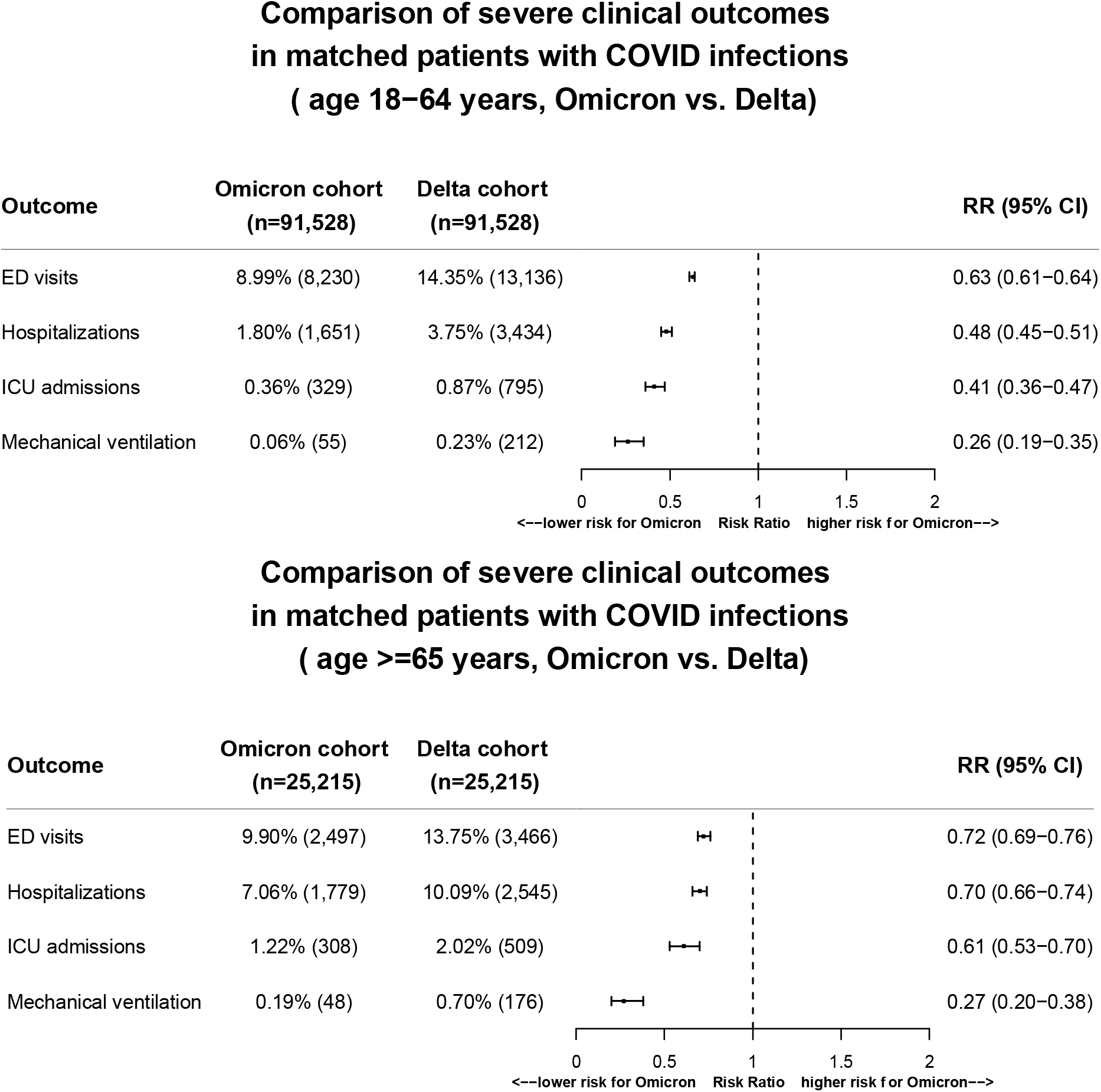
Age-stratified comparison of severe clinical outcomes in the 3-day time-window that followed from the first day of SARS-CoV-2 infection between Omicron and Delta cohorts that were propensity-score matched for demographics (actual age at the time of COVID infection, gender, race, ethnicity), socioeconomic factors, COVID-19-reated health conditions, medications, and documented vaccination status.

### Racial and ethnic disparities in severe clinical outcomes for the Delta and Omicron cohorts

There was significant difference in ED visits in the 3-day time-window after COVID infections between infected Black and White patients after propensity-score matching for age, gender, socioeconomic factors, morbidities, medications, and documented vaccination status. In the Omicron cohort, the rate for ED visits in Black patients was 14.4%, higher than 7.8% in the matched White patients (RR: 1.83 [1.75-1.92]). Similar racial disparity in ED visits was for the Delta and Delta-2 cohorts (**Figure 4a**). Significant difference in ED visits were observed between matched Hispanic and non-Hispanic patients in the Omicron, Delta and Delta-2 cohorts. In the Omicron cohort, the rate for ED visits in Hispanic patients was 25.2%, higher than 12.8% in the matched non-Hispanic patients (RR: 1.98 [1.85-2.11]) (**Figure 4a**). Hospitalization risk did not differ between matched Black and White patients in the Omicron cohort but was higher in Black patients than White patients in the Delta cohort (RR: 1.22 [1.17-1.26]) and Delta-2 cohorts (RR: 1.23 [1.14-1.34]) (**Figure 4b**). The overall lower risks for hospitalizations and smaller sample sizes in the Omicron could accounted for the trend level difference between Black and White patients. No marked difference in hospitalizations was observed between Hispanic and non-Hispanic patients after matching for age, gender, socioeconomic factors, morbidities, medications, and documented vaccination status (**Figure 4b**). There was significant difference in ICU admissions between matched Black and White patients in the Omicron, Delta and Delta cohorts (**Figure 4c**). In the Omicron cohort, the risk for ICU admissions in Black patients was 0.43%, higher than 0.28% in the matched White patients (RR: 1.54 [1.17-2.01]). Similar difference in ICU admissions were observed between matched Hispanic and non-Hispanic patients. In the Omicron cohort, the risk for ICU admissions in Hispanic patients was 0.75%, higher than 0.51% in the matched non-Hispanic patients (RR: 1.48 [1.01-2.16]).

**Figure 4.**
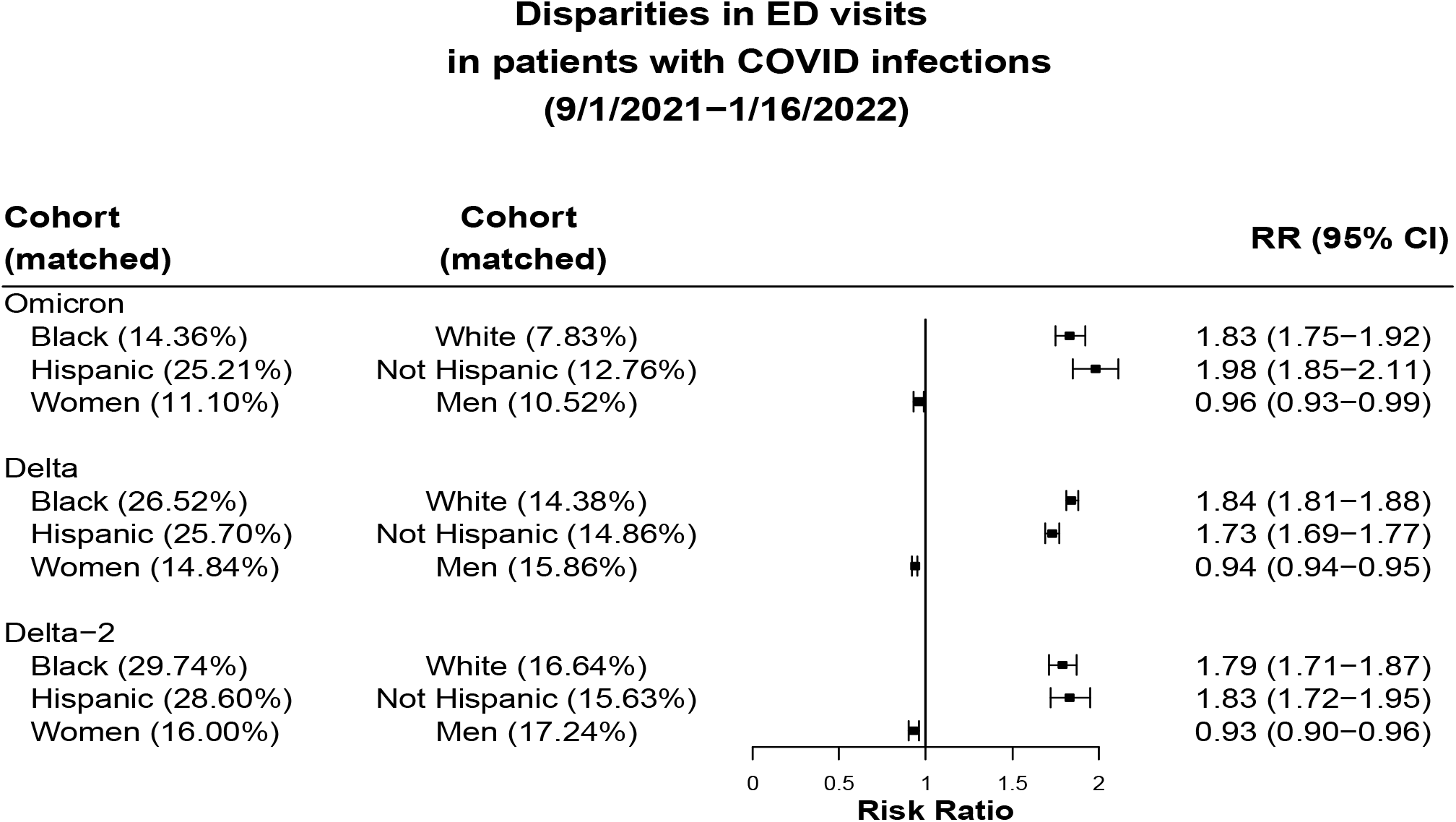
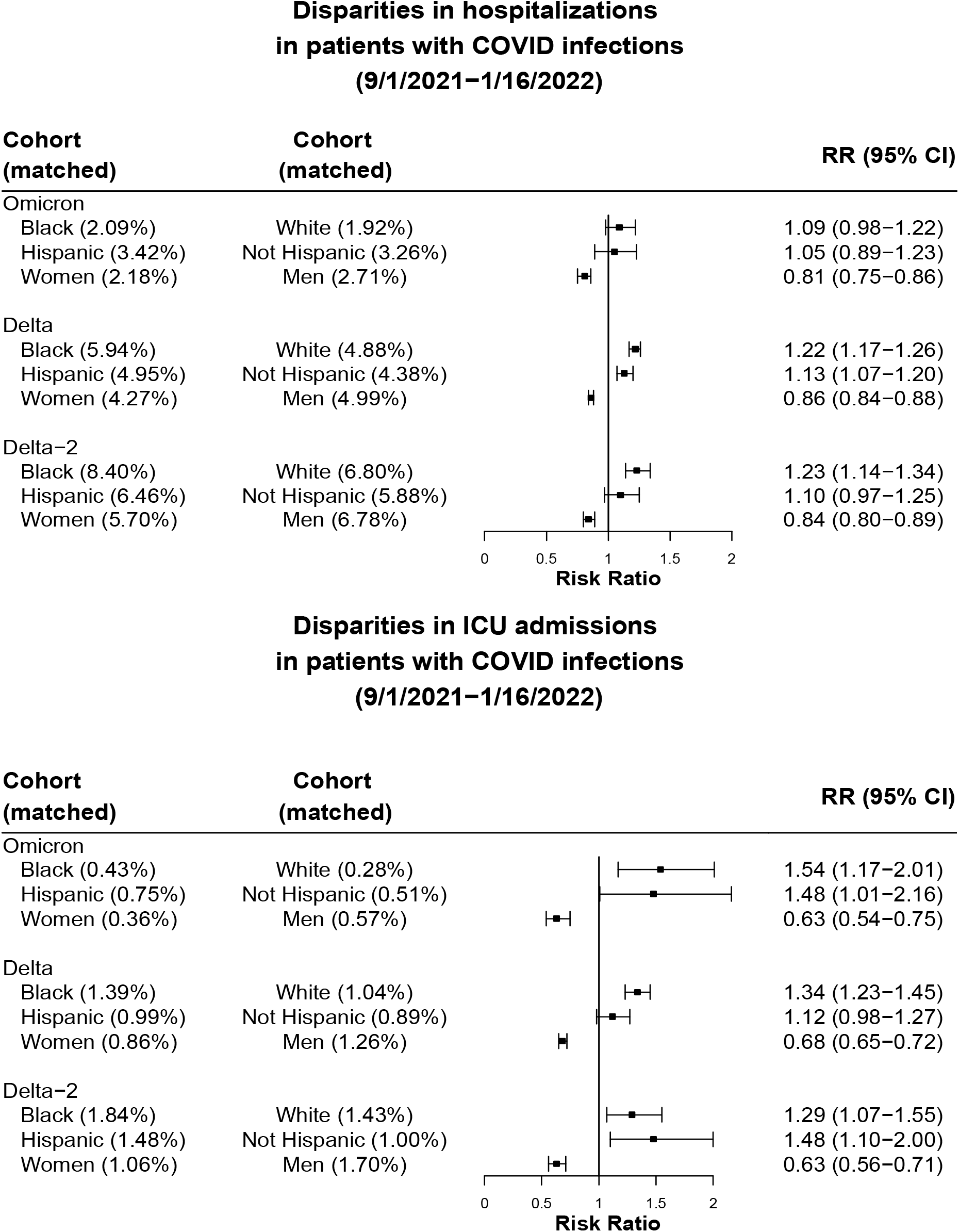
Comparison of ED visits, hospitalization, ICU admission in the 3-day time-window that followed from the first day of SARS-CoV-2 infection between matched Black vs White patients, Hispanic vs non-Hispanic patients, women vs men in the Omicron, Delta, and Delta-2 cohorts, respectively. Race, ethnicity, or gender stratified cohorts were propensity-score matched for other demographics, socioeconomic factors, COVID-19-reated health conditions, medications, and documented vaccination status.

Women had lower risks than men for ED visits, hospitalizations and ICU admissions in the Omicron, Delta and Delta-2 cohorts, after matching for race, ethnicity, socioeconomic factors, morbidities, medications, and documented vaccination status. This gender difference was especially profound for ICU admissions in all cohorts. In the Omicron cohort, the risk for ICU admissions in women was 0.35%, compared to 0.57% in men (RR:0.63 [0.54-0.75]) (**Figure 4c**).

## Discussion

This study, using a nation-wide database of EHRs in the US, showed that the monthly COVID infection rate among patients who had no prior infections was stable between September and November 2021 when Delta predominated. Starting December 2021, the infection rate increased, coincident with the emergence of the more transmissible Omicron variant and reached 6 times that of the earliest time period by the second half of December, when Omicron predominated. These results are consistent with Omicron being more infectious than the original SARS-CoV-2 virus(10) and provide new evidence that Omicron is more infectious and spreads more easily compared to Delta. The infection rate in January 2022 (1/1-1/20) was 5.2 new cases per 1000 persons per day, higher than 3.8 in second half of December 2021, indicating that infections with Omicron had not yet plateaued. Children under 5 years had the highest incidence rate of COVID infection among the four age groups during both Delta and Omicron time, which could reflect lower immunity due to their ineligibility for COVID-19 vaccines. Overall infection rates decreased with increasing age, which correlated with the higher vaccination and booster rates among older age groups. Accordingly, patients over 65 years old had the lowest infection rate, which may reflect high vaccination or booster rates and more preventive measure adoptions among older adults. Close monitoring of infection rates among all age groups are necessary given the potential for further viral mutations and emergence of other variants.

Significant racial and ethnic disparities in COVID infection rates were observed, with highest rates in Black patients, followed by Hispanic patients across all age groups. The disparities further widened during the Omicron time. During the Delta period, the infection rate in Black patients was 1.3-1.4 times of that in White patients, and the incidence rate in Hispanic patients was 1.6-1.8 times of that in non-Hispanic patients. During the Omicron period, the incidence rate in Black patients was 3-4 times of that in White patients, and the incidence rate in Hispanic patients was 3 times of that in non-Hispanic patients. Studies showed that Black patients had higher risks for COVID infection compared to White patients in the early stages of the pandemic when vaccines were not available(11–16). However among the fully vaccinated patients, the risk for breakthrough COVID-19 infection did not differ between Black and White patients(17). Reasons underlying the profound racial and ethnic disparity in the COVID infection rates during the Omicron predominance period need to be examined, which could include differential vaccination uptake by racial and ethnic groups, living situations, occupations that might expose them to greater risk of infections among others.

SARS-CoV-2 infected patients in the Omicron cohort were demographically different from and had fewer adverse health conditions than in the Delta cohort. After matching for age, gender, race/ethnicity, adverse socio-economic determinants of health, COVID-related comorbidities and medications, and EHR-documented vaccination status, the risks for severe clinical outcomes in the 3-day time-window following initial SARS-CoV-2 infection in the Omicron cohort were significantly lower than in Delta cohort, with 30% reduction for ED visits, 42% reduction for hospitalizations, 53% reduction for ICU admissions and 75% reduction for mechanical ventilation use, even when after matching the Omicron cohort had more comorbidities than the Delta cohort. Propensity score matching indicates that the milder disease severity for the Omicron cohort compared to the Delta cohort was not confounded by age, race/ethnicity, overall health of those infected, socioeconomic factors, medications, and vaccination status. Control studies comparing the matched Delta-2 and Delta cohorts showed no such reductions in disease severity, suggesting that the findings were not confounded by other factors such as timing, colder weather, holiday seasons, and more vaccination and boosters for later times. Similarly, significantly lower disease severity was observed for Omicron compared to Delta in children under 5 years old. Since vaccines are not available to children under 5 years, there were no confounding effects from vaccination. In addition, the study population comprised patients who had no prior COVID-19 infection, therefore confounding effects from previous infections were minimized. Taken together, these results suggest that the differences in disease severity between the Omicron cohort and the Delta cohort resulted from inherent properties of the variant itself. Future studies are needed to examine how vaccination and pre-existing immunity from prior infections further alter the clinical course and outcomes of COVID infections with Omicron.

There were significant racial and ethnic differences in ED visits in 3-day time-window that followed from the first day of SARS-CoV-2 infection in both the Omicron and Delta cohorts, with higher rates in Black and Hispanic patients compared with White and Non-Hispanic patients respectively. A previous study reported racial and ethnic disparities in ED visits for COVID-19 between October and December 2020, with Black and Hispanic persons experienced 1.4-1.7 times the rate of ED visits than White persons(18). Our study showed that these racial and ethnic disparities in ED visits persisted and widened during the Delta and Omicron predominance periods, with Black patients experincing 1.8 times the rate of ED visits than White patients, and Hispanic patients experiencing 2.0 times the rate of ED visits than non-Hispanic patients, even after propensity-score matching for other demographics, comorbidities, socioeconomic factors, morbidities, medications, and documented vaccination status. The reasons underlying this persist racial and ethnic disparity in ED visits associated with COVID-19 warrant further investigation, including racial/ethnic inequity in health status and disease outcomes, lack of primary care in health disparity groups

Racial and ethnic disparities for COVID-related hospitalizations and mortality were observed in the early stages of the pandemic before vaccines were available(11). Findings from this study show significant racial and ethnic differences in ICU admissions in 3-day time-window that followed from the first day of SARS-CoV-2 infection. During the Omicron predominance period, Black patients experienced 1.5 times the rate of ICU admissions than White patients, and Hispanic patients experienced 1.48 times the rate of ICU admissions than non-Hispanic patients, after propensity-score matching for other demographics, comorbidities, socioeconomic factors, morbidities, medications, and documented vaccination status. Potential explanations given for these disparities include disproportionate burden of health conditions, socioeconomic status, genetic or other biological factors, community characteristics, air pollution, among others. A recent study showed racial and ethnic disparities in receipt of medications for treatment of COVID-19, with lower use of monoclonal antibody, remdesivir and dexamethasone among Black and Hispanic patients than White and Non-Hispanic patients(19). Though our study adjusted for comorbidities, socioeconomic status, behavioral factors including smoking and alcohol drinking, COVID medication usage and documented vaccination status, certain residual and unmeasured confounding factors (e.g., life-style factors, neighborhood, community living, genetics) may have contributed to the observed racial and ethnic disparities in COVID-19 outcomes. This study showed marked racial and ethnic disparities in short-term clinical outcomes of COVID-infections. Studies are needed to investigate the longer-term effects of COVID-19 on health disparities and to understand risk factors for the differential outcomes in disproportionately affected racial/ethnic groups.

Research and clinical data on gender disparities in COVID-19 infection and severe clinical outcomes are limited(20,21). Early pandemic data from Europe and China showed that COVID-19 was associated with more death in men than in women(22,23). Findings from our study showed that the COVID infection rates were consistently lower in women than in men during both the Delta and Omicron periods. In addition, women had lower risks for ED visits, hospitalizations and ICU admissions compared to men, after matching for race, ethnicity, socioeconomic factors, morbidities, medications, and documented vaccination status. A prior study showed that women had a lower likelihood to be admitted to ICU than men regardless of disease severity, despite being more severely ill(24). Our study shows that in the context of COVID-19, women consistently reported lower infection rates and experienced fewer ED visits, hospitalizations and ICU admissions. Future studies are needed to disentangle the impacts of gender-specific factors such as health status and biological factors, healthcare usage, lifestyle, psychological stress, socioeconomic conditions, and treatments.

Our study has several limitations: First, the observational, retrospective nature of this study of patient EHR data could introduce case selection biases, over representation of symptomatic testing, reporting and follow up issues. However, because we compared the different cohort populations all from the TriNetX dataset, these issues should not substantially affect the relative risk analyses. Second, patients in the TriNetX EHR database are those who had medical encounters with healthcare systems contributing to the TriNetX Platform and do not necessarily represent the entire US population. Therefore, results from the TriNetX platform need to be validated in other populations. Third, both the Omicron and Delta cohorts in our study were defined based on CDC’s national genomic sequence surveillance and not by variant identification in individual patients. Therefore, the Omicron cohort likely contained a few infections with the Delta variant. However, our findings of reduced disease severity in the Omicron cohort compared to the Delta cohort are consistent with findings from Africa(2), Scotland(3), and England(4) that were based on individual genomic sequences. The fact that no reduction in severe disease frequency was observed between the two Delta cohorts further suggest that the differences resulted from inherent properties of the variant itself. Moreover the increase in severity outcomes for the Delta-2 compared to the Delta cohort, if anything, points to waning immunity from vaccination during this time period, though this is confounded by uptick in boosters(25). Fourth, the vaccination status of the infected individuals in TriNetX database is incomplete. The vaccination rate documented in TriNetX is only about 8-14%, whereas the reported vaccination rates during this time period(26) indicate that most patients in the study population were likely to have been vaccinated. The incompleteness of vaccination status information may have not significantly affect the relative risk analyses, as the percentages of vaccination among propensity-score matched Omicron and Delta cohorts were similar and cohorts were matched for demographics, adverse socioeconomic determinants of health and comorbidities, many of which are associated with vaccination acceptance(27,28). However we were unable to examine how vaccination impacted disease severity of Omicron. Finally, the findings apply only to infections that occurred in the US between 12/26/2021– 1/16/2022 when Omicron accounted for more than 92% of all variants. Given the potential for further viral mutations, future studies are needed to closely follow up disease severity as well as longer term outcomes associated with infections from Omicron or other variants.

In conclusion, the incidence rate of COVID infection during the omicron predominant period (prevalence >92%) was 6-8 times higher than during the Delta predominant period that preceded it consistent with greater infectivity. The incidence rate was highest among those less than 5 years of age, Black patients and Hispanic patients. COVID infections occurring when the Omicron predominated were associated with significantly less frequent severe outcomes than in matched patients when the Delta variant predominated. There were significant racial, ethnic and gender disparities in severe clinical outcomes, with Black and Hispanic patients and men disproportionally impacted. Though the lower severity observed in the Omicron cohort is encouraging, further studies are needed to monitor longer-term acute consequences from Omicron infection, especially mortality, the propensity for development of “long COVID”, the rapidity of spread, potential for mutation, how prior infections and vaccination alter clinical responses and to characterize the factors that underly the vulnerability for severe outcomes in Blacks and Hispanics. Additionally, although severe infections from the Omicron variant, based on this analysis, appear to be less frequent, because of Omicron’s increased transmissibility, many more individuals have been infected, so the overall number of ED visits, hospitalizations, ICU admissions, and mechanical ventilator use in infected people may still be greater with the Omicron variant than the Delta variant.

## Data Availability

All data produced in the present work are contained in the manuscript

## Contributors

RX conceived, designed, and supervised the study, performed literature review and reviewed and drafted the manuscript. LW conducted all the experiments, performed data analysis and prepared tables and figures. NAB, PBD, DCK, and NDV critically contributed to study design, result interpretation and manuscript preparation. We confirm the originality of content.

## Acknowledgments

We acknowledge support from National Institute on Aging (grants nos. AG057557, AG061388, AG062272), National Institute on Alcohol Abuse and Alcoholism (grant no. R01AA029831), National Institute on Drug Abuse (grant no. UG1DA049435), the Clinical and Translational Science Collaborative (CTSC) of Cleveland (grant no. 1UL1TR002548-01), National Cancer Institute Case Comprehensive Cancer Center (R25CA221718, P30 CA043703, P20 CA2332216).

## Meeting Presentation

No

